# TEXT MINING OF THE PEOPLE’S PHARMACY RADIO SHOW TRANSCRIPTS CAN IDENTIFY NOVEL DRUG REPURPOSING HYPOTHESES

**DOI:** 10.1101/2022.02.02.22270107

**Authors:** Rahul Yedida, Jon-Michael Beasley, Daniel Korn, Saad Mohammad Abrar, Cleber C. Melo-Filho, Eugene Muratov, Joe Graedon, Terry Graedon, Rada Chirkova, Alexander Tropsha

**Author notes:** Corresponding Authors: Rada Chirkova, Department of Computer Science, North Carolina State University, Raleigh, North Carolina, USA; Alexander Tropsha, UNC Eshelman School of Pharmacy, University of North Carolina at Chapel Hill, Chapel Hill, North Carolina, USA.

## Abstract

**Objective:** Social media mining may provide surprising information about unknown effects of drugs. We endeavored to uncover such unknown drug-disease relationships by text mining of audio record transcripts from the popular NPR show, The People’s Pharmacy.

**Materials and Methods:** We used Google Cloud to transcribe episodes of the NPR podcast into textual documents. We then built a pipeline for systematically pre-processing the text to ensure quality input to the core classification model. Finally, results of the model were filtered by a series of post-processing steps. Our classification model itself uses the FLAIR language model pre-trained on PubMed abstracts. The modular nature of our pipeline allows for ease of future developments in this area by substituting higher quality components at each stage of the pipeline. To validate the drug-disease relating assertions extracted from the podcast, we utilized the DrugCentral database and ROBOKOP biomedical knowledge graph, which capture drug-disease relationships for FDA approved medications.

**Results:** Our model identified 128 drug-disease pairs that were found in DrugCentral and 112 novel candidate pairs requiring expert review. To demonstrate the expert review process, we found literature evidence supporting the assertions for novel drug-disease pairs.

**Discussion and Conclusion:** Text mining of social media is increasingly used to uncover novel relationships between semantic concepts corresponding to biomedical concepts. However, mining audio transcripts of specialized podcast shows has not been explored previously for this purpose. Using this approach, we have identified several unknown drug-disease relationships with support in biomedical literature. The proposed approach can extend beyond radio podcasts and could be applied to any source of audio and textual data.

## 1. BACKGROUND AND SIGNIFICANCE

Major computational approaches to drug discovery include so called ligand-based (such as similarity search, pharmacophore modeling, Quantitative Structure – Activity Relationship (QSAR) modeling) and structure-based (docking and virtual screening) approaches [1,2]. These approaches have been extensively employed to discover drugs against multiple diseases including recent gargantuan efforts to repurpose drugs against COVID-19 (reviewed recently by Muratov et al. [3]). In recent years, literature-based discovery (LBD) has been added to the arsenal of tools employed specially to identify novel or poorly known uses of existing drugs (also known as ‘*drug repurposing*’ [4]) or identify potential side effects of the existing drugs [5]. Commonly, LBD approaches have been used to analyze textual sources such as abstracts of full texts of the scientific papers [6,7]) although in some cases social media mining has been explored as well [8]. In this study, we proposed an uncommon LBD approach based on text mining of <u>transcribed</u> audio podcast recordings. Herein, we describe a systematic, step-by-step approach for extracting meaningful, nontrivial pairs of drug-disease concepts from raw audio files that are transcribed to unstructured textual information sources.

There have been previous attempts to leverage nontraditional data sources in the medical community. For instance, Mohammadhassanzadeh et al. [9] and Noh et al. [10] used podcasts as knowledge sources. The freeform communication typical of podcasts produces a substantial amount of natural language data, which can be mined for useful information. Many of these podcasts take the form of a dialogue between a host or hosts and guest, these guests may or may not be medically trained. Because of their freeform nature, podcasts contain language with little structure to be exploited (when compared to an academic paper), with episodes taking the form of dialogues or unscripted interviews. Mining such unstructured text is a fundamentally challenging Natural Language Processing (NLP) task [11]. Recent advances in NLP systems have enabled the training of so-called “language models”, multi-task general-purpose learners that model the structure of natural language sentences and perform various tasks such as part-of-speech tagging [12]. The most famous of these language models is the recent BERT model by Google [13], which achieved state-of-the-art results on several NLP tasks. However, we choose to use a different state-of-the-art language model, FLAIR [14], for the modular nature of its code and its ease of use. We built our code upon the recent work of BioFLAIR [15], a model that fine-tuned FLAIR on PubMed data. By using this model to mine transcribed episodes of the popular NPR radio show ‘People’s Pharmacy”, we have identified over one-hundred known drug-disease connections (model validation) as well as over a hundred of connections not annotated in popular drug registries such as DrugCentral [16].

In summary, we have developed and validated a novel framework for extracting meaningful drug-disease pairs from unstructured, natural language. This framework is built on a modular *plug-and-play* system that can be easily modified to include new algorithms or data sources. We have emphasized the importance of rigorous curation of primary data and validation of the assertions generated by this approach. The code and software developed in this project have been made publicly accessible at https://github.com/yrahul3910/drug-repurposing-textmining.

## 2. DATA AND METHODS

Our approach comprises three major parts: pre-processing, pair formation, and post-processing. Key steps of our pipeline are shown in Figure 1.

**Figure 1.**
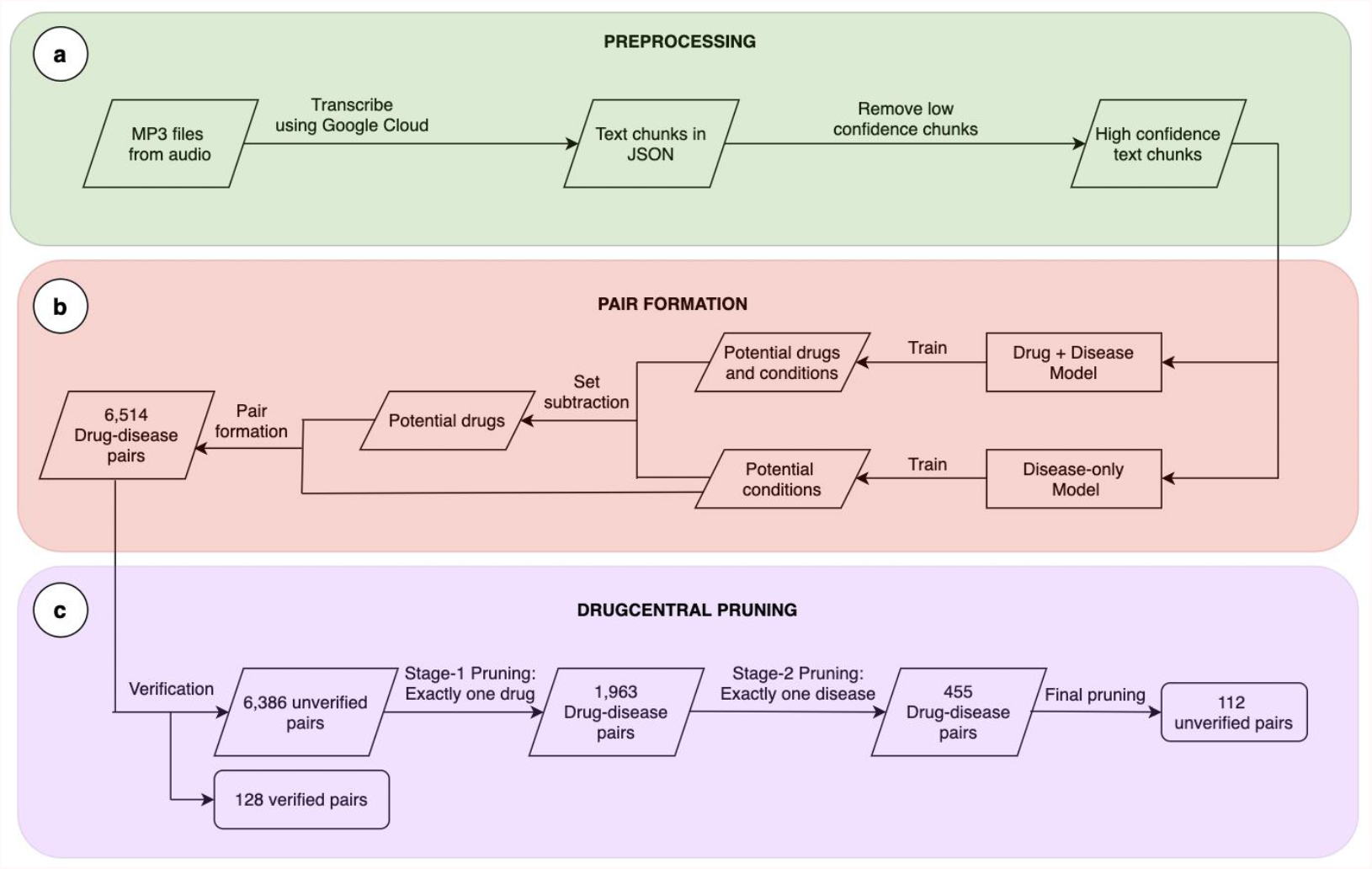
The workflow for text mining of the People’s Pharmacy episodes. The workflow is comprised of three components: a) Creation of textual data from podcasts. b) The process of mining drug-disease pairs from transcripts of the podcast. c) Verification of drug-disease pairs using DrugCentral.

### 2.1 The source dataset

The People’s Pharmacy NPR Radio Program is a popular weekly radio program hosted by Joe and Terry Graedon. This show has been running for forty years, with over one thousand episodes having been recorded. Episodes of The People’s Pharmacy focus on the intersection of the general public and the medical field, cover topics such as: the effectiveness of drugs, home remedies, and doctor-patient interactions [17].

The input podcast data included all available MP3 files for The People’s Pharmacy, 199 episodes in total [18]. The raw MP3 files were passed through Google Cloud Text-to-Speech (TTS)^1^ to transcribe the voice recordings to text. The result of the Google Cloud transcriptions are JSON files for each episode. These JSON files are chunked by Google (into arbitrary sizes). Each chunk was given a TTS confidence score by the transcription. On average, each episode had 81 chunks. Figure 2 illustrates the distribution of the number of text chunks in the episodes. In total, we processed 16,291 chunks.

**Figure 2.**
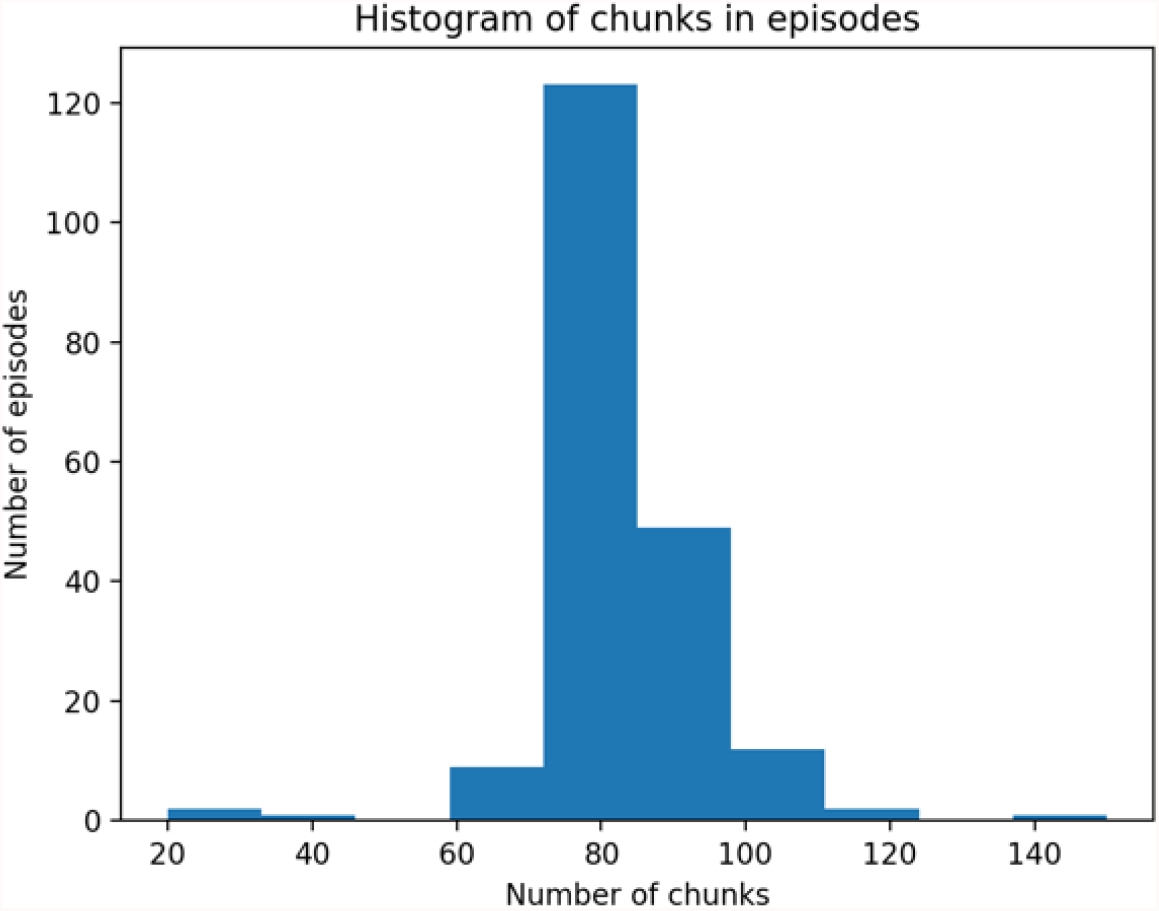
A histogram of the number of chunks of text produced per podcast episode. These chunks are generated automatically by Google Cloud’s transcription service. Here we show how in general each episode produces about the same amount of content.

### 2.2 Auxiliary datasets

Our approach relies on training models in an unsupervised manner on large-scale datasets *a priori*. This is done based on the domain of the problem—in this case, drugs and diseases. Specifically, we use the *BioCreative V Chemical Disease Relation* (*bc5cdr*) corpus [19] and the NCBI disease corpus [20], which contains 793 PubMed abstracts fully annotated. We choose these because a prior implementation that fine-tuned word embeddings for medical corpuses, BioFLAIR[15], used a superset of these datasets. However, for the purposes of our approach, these two corpuses were sufficient. These datasets were obtained from the BioFLAIR [15] code repository.

### 2.3 Pre-processing

Our first pre-processing step (performed on the TTS transcription output) was removing chunks that have a confidence score less than 96%. We derived this threshold by manually going through 100 chunks of text and analyzing poorly transcribed chunks of text (as a particularly egregious example, “Dr. Sacks” was transcribed as “Doctor Sex”). We found that chunks with a confidence score less than 96% contained errors which would reduce our ability to find valid medical information.

Because our core idea was to identify related drug and disease terms (together called “medical terms”), we sought a language model to identify these medical terms. However, traditional language model would also identify “medical-adjacent” terms, such as “doctor” and “hospital”. It is not informative to learn that visiting a doctor helped cure a patient of some disease, so next we compiled a list of such medical-adjacent terms. During our pass through 100 random chunks of text, we were able to compile a list of these terms. This compiled list is summarized in Table 1. We performed a partial string match and removed all medical-adjacent words from all chunks of text.

**Table 1:** List of medical-adjacent terms

|  |  |  |
| --- | --- | --- |
| pharmacy | health | allergist |
| patient | biochemical | medication |
| symptom | poop<sup>2</sup> | feces |
| urologist | polypharmacy | relief |
| nutrient | dr | herbal |
| care/healthcare | prognosis | diagnosis |
| die/death | insurance | remittance |
| relapse | physician | decease |
| practitioner | anesthesia | ICU |
| Medicare | etiology |  |

### 2.4 Classification

The kind of problem we described can be framed as a Named Entity Recognition (NER) task. Specifically, we endeavored to classify each word in the text as a drug, a disease, or neither. To do so, a language model was used. Such models can be adapted to several tasks because of the general-purpose nature of their initial training. As shown in Figure 3, given a sentence “George Washington was born”, an NER model would be expected to predict that “George” is the beginning of a *PERSON* entity, and “Washington” was the corresponding ending. All other words are assigned an *OTHER* entity. In our case, the model predicts *B-DRUG* and *E-DRUG* to delimit drug candidates, and *B-DISEASE* and *E-DISEASE* to delimit disease candidates (note that at the time of prediction, whether a candidate is truly a drug/disease is unknown). A BiLSTM-based model [21,22] is used to achieve this, which is implemented by the FLAIR framework [23]. The overall architecture of the model is illustrated in Figure 3.

**Figure 3.**
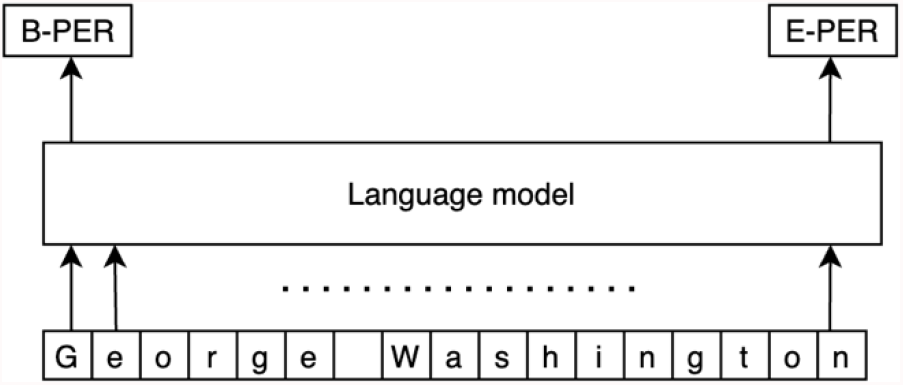
A demonstration of how a language model from FLAIR would process the name, “George Washington”, using B-PER (beginning of person) and E-PER (end of person) outputs.

Because of the high modularity and accuracy of the FLAIR language model in state-of-the-art datasets, we leveraged it to train a model that fits our task. At a high level, we used the embeddings trained on PubMed, and pre-train it on our data. Finally, we ran NER on each chunk separately.

In detail, we used two instances of the FLAIR model [23]: We fine-tuned one model on the bc5cdr data [19], which contain labels for identifying both diseases and drugs, and fine-tuned the other model on the NCBI disease dataset [20], which contains only disease labels. Next, we identified the drugs in text as a set difference between the two models’ outputs. We used this twin classification model because it leads to more stable results. Specifically, this approach has two advantages: (1) using this model yields less pairs for a human expert to verify; (2) for a pair to be proposed, it needs to be acknowledged by two differently trained models; this is akin to asking for two different perspectives from two experts, each of whom studied the same material from different sources.

This robust approach is not very computationally expensive. In our testing, the significant computation (besides the fine-tuning) was in initializing the models, not in classifying. Therefore, our approach is fast, scalable, and robust in identifying perplexing drug-disease pairs for further examination from a human expert.

### 2.5 Post-processing

Our post-processing steps are designed to work in conjunction with the classification step and prune the pairs proposed. We disregarded adjacent medical terms of the same class (i.e., a drug cannot treat another drug; a disease cannot treat another disease), so that the final proposed pairs were at least semantically valid. We also removed adjacent duplicate medical terms. Finally, we, removed duplicate pairs (including pairs of the form A, B and B, A). As shown in Figure 1, we began with the original 6,514 drug-disease pairs proposed and verified 128 pairs as exact matches in DrugCentral; the 6,386 remaining pairs were sequentially pruned using the DrugCentral database by applying the following sanity checks in order: (a) Exactly one of the terms of the pair must be a drug—this led to 1,963 pairs. (b) Exactly one of the terms must be a disease—this led to 455 pairs. (c) Both (a) and (b) must be true simultaneously, i.e., exactly one of the terms must be a drug and exactly one of the terms must be a disease—this led to the final 112 pairs that were shown to a human expert for verification.

## 3. RESULTS

Amongst the 16,291 text chunks in a total of 199 episodes, 6,514 pairs of drug-disease associations were found by our model, which were pruned to 240 pairs. These pairs were then cross-referenced against the DrugCentral database, with 128 pairs matching against known facts and being verified, and 112 pairs that needed further review. This gives us a precision score of 128/240 = 53.3%. An example of using this approach for a sample text chunk is shown in Figure 4. As illustrated, the pair formation module creates associations between the adjacent medical terms. Then, set subtraction was used to extract only drug-disease pairs from the raw data.

**Figure 4.**
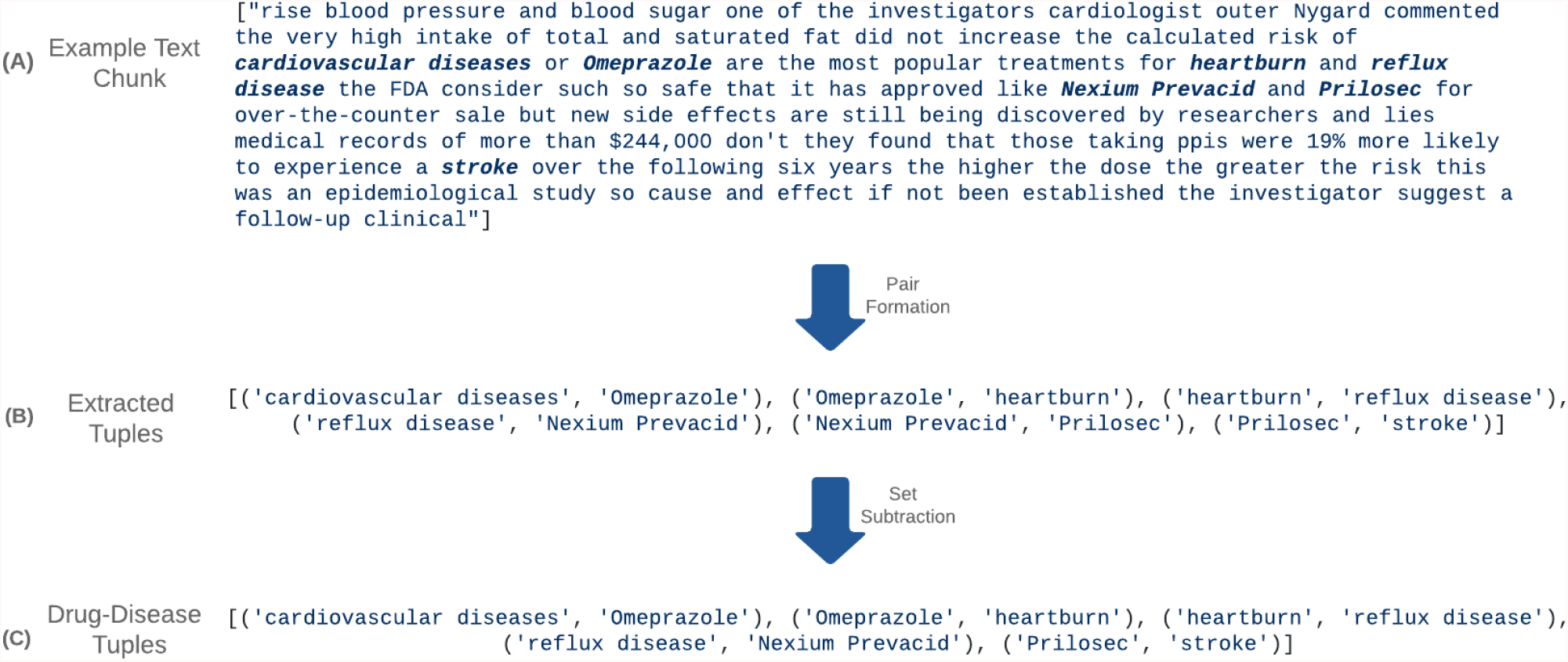
Drug-Disease tuple generation from raw text chunks: In this example we see the portions of the text where pairs appear in (A) bolded. These highlighted sections are then reduced to pairs of biomedical terms in (B). These pairs are then reduced further to exclusively drug/disease tuples in (C).

Due to several text chunks reporting the same kind of disease associations, several duplicate pairs were reported. Hence, deduplication was performed as a post-processing step.

The results indicate that the model was able to find known pairs of drug-disease associations, but it also yielded 112 unverified associations. We used ROBOKOP, a biomedical knowledge graph[24], to formulate plausible connections between the proposed pairs, then conducted a manual literature review to fill in additional details of these connections. In the following section, we discuss some associations that we confirmed through this process. ROBOKOP is a knowledge graph constructed from forty-five independent biomedical databases, it aims to integrate all of these disparate sources of information into a singular platform, where the data may be interoperable and queried through a single interface. Biomedical concepts (such as a disease like *Chordoma*, or a drug like *aspirin*) will be represented as nodes in the graph, how these ideas relate to each other is represented as edges connecting these nodes.

### 3.1 Examples of associations found in the transcripts and confirmed by ROBOKOP

The list of associated drug-diseases terms contained some pairs that could be less obvious or less studied than others and could become interesting new hypotheses to guide research. For example, an association between asthma and fluticasone that is validated by ROBOKOP would be obvious to a pharmacologist studying drug repurposing because fluticasone is officially indicated for the management of asthma symptoms [25]. To demonstrate our method’s ability to identify novel associations, we used the DrugCentral Online Drug Compendium (https://drugcentral.org/) to distinguish “obvious” and “non-obvious” drug-disease associations. Drug-disease associations not listed as official indications in DrugCentral were pruned from the list and 112 non-obvious associations emerged.

To demonstrate the validation of these non-obvious pruned pairs, we employed ROBOKOP to derive clinical outcome pathways (**COP**). A COP is an emerging concept [26] designed to be more informative than mechanism of action because it describes the cause-and-effect chain of chemical-biological interactions underlying clinical effects of drugs [26,27]. ROBOKOP is well-suited for helping an expert elucidate a COP by identifying intermediary biological entities (genes, cell types, biological processes) involved in a drug’s clinical effect. For the non-obvious pruned pairs, we used ROBOKOP to identify drug--gene--disease connections to serve as the starting point for manual literature review by an expert. This method helps saves time and expense of using a human to explore novel drug-disease associations. Two examples of COPs elucidated by using ROBOKOP are presented below and illustrated by Figures 5 and 6.

**Figure 5.**
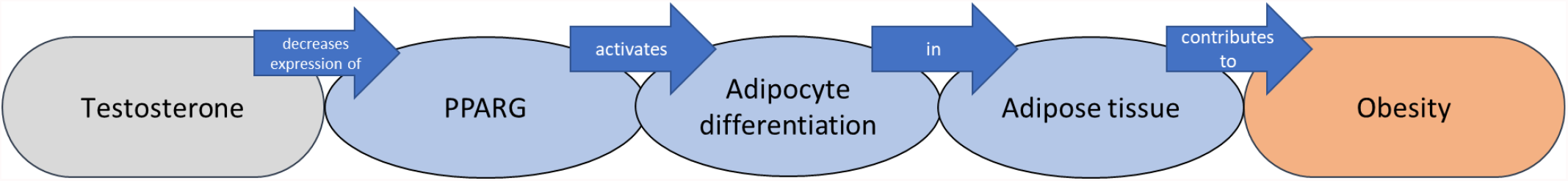
Testosterone to obesity COP elucidated through manual literature using PPARG gene identified from ROBOKOP as literature search inspiration.

**Figure 6.**
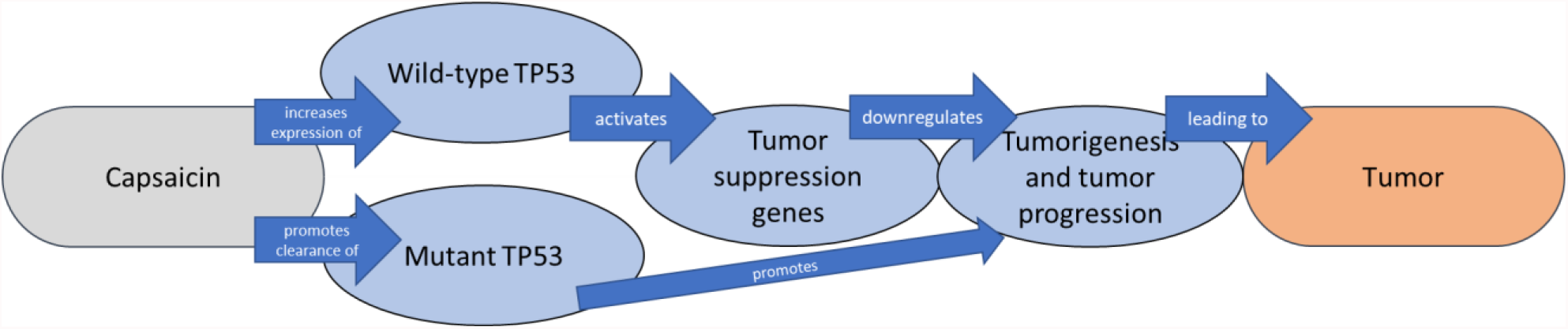
ROBOKOP identified TP53 as gene intermediate for capsaicin-tumor COP. Literature review elucidated capsaicin’s action on both wild-type and mutant TP53 isoforms.

#### 3.2.1 Obesity and Testosterone

Testosterone (T) deficiency has been linked to male obesity in a bidirectional manner, with obesity contributing to low T, and low T likewise contributing to obesity [28]. T decreases the expression of peroxisome proliferator activated receptor gamma (PPARG) in ovarian granulosa cells [29]. Additionally, the presence of dihydrotestosterone (DHT) results in reduced activity of PPARG in fat-storing adipocytes [30]. PPARG is a nuclear receptor and transcriptional cofactor for genes controlling fat storage and metabolism in adipose tissue [31]. This evidence taken together suggests a potential COP for the treatment of obesity with T supplementation and supports the testosterone – obesity association.

#### 3.2.2 Tumors and Capsaicin

Capsaicin, the “spicy” ingredient in chili peppers, promotes the degradation of mutant tumor protein p53 (TP53) and promotes the reactivation of wild-type TP53 function [32]. TP53 is a transcription factor protein important to cancer prevention and response to acute stressors [33]. Tumor-promoting mutations in TP53 have been observed in most human cancer types, especially ovarian, colorectal, esophageal, head and neck, and laryngeal cancers with frequencies of 40-50% [34]. Capsaicin induces autophagy and clearance of mutant TP53 while promoting DNA repair to restore wild-type function [35]. The mechanism by which a molecule like capsaicin can a disease becomes clear when all of these molecular interactions are taken together and arranged in the form of a COP. These capsaicin-induced molecular events could facilitate tumor cell death and represent promising COPs for capsaicin’s reported anti-tumor properties.

## 4. DISCUSSION

We discuss some of the aspects and potential concerns with our method and seek to provide additional reasoning for our approach in this section.

### 4.1 Text chunking

It may be noted that eliminating some of the chunks adds discontinuity to the text. However, we argue that this should not significantly impact our results; rather, it helps generate more robust results. By eliminating chunks below the threshold, we start with better, more robust data. We argue that this makes it less of a burden on the rest of the process to filter out bad results.

Furthermore, by not running our model on two continuous chunks, we avoid erroneous results generated by mixing two potentially discontinuous chunks of text as input.

### 4.2 Irrelevant term removal

Simply removing the irrelevant terms from the text may cause the resulting text to become ungrammatical and incoherent. However, because our approach does not have a strong reliance on the structure of the sentence, this does not affect our results. Specifically, our approach is based on adjacent medical term pair formation; by removing potentially misleading terms, which we call medical-adjacent terms, we remove potential bad pairs and obtain more robust results.

### 4.3 Pair formation

Because the BioFLAIR classifier identifies both diseases and treatments as medical terms, it is certainly possible to have pairs formed consisting of only diseases or only treatments. To alleviate this, we used a twin classifier model in the classification and pair formation stage. We train one BioFLAIR model as described above, and another that identifies only diseases. Then, we form pairs only from the set difference of the terms detected by these models. We find this significantly reduces (by 64.5%) the number of pairs suggested by the overall pipeline and improves the overall quality of the results.

### 4.4 Validation

The validation step has been a huge concern given the prohibitive number of pairs generated to be manually checked. Although, ROBOKOP provided a platform for validation of possible associations, some true positives given by the model are not present in the underlying ROBOKOP knowledge graph. Additionally, there were also some false positives found in the ROBOKOP Knowledge Graph, where trivial relationships that are found by the model (but do not represent meaningful disease-treatment pairs) are marked as correct. This highlights the importance of confirming ROBOKOP COP pathways through manual literature review, as performed in the two case studies shown.

## 5. CONCLUSION

This study aimed to construct an automated pipeline to identify drug-disease tuples from unstructured texts using The People’s Pharmacy transcripts as an experimental dataset. Our current pipeline achieves a 53.3% precision score on the DrugCentral database. To the best of our knowledge, our approach is the first end-to-end, modular, automated system to propose novel drug-disease associations from unstructured verbal records. Our precision score indicates that around half the proposed pairs are already known to be true. Further, our multiple levels of pruning ensured that only 240/6514 = 3.7% of the total pairs have required human evaluation.

In summary, we used a state-of-the-art NLP system as part of a pipeline to identify plausible novel treatments for diseases from unstructured text. These hypotheses can be validated by human experts, who may decide which are worth pursuing further.

## Data Availability

The code and software developed in this project have been made publicly accessible at https://github.com/yrahul3910/drug-repurposing-textmining.

## Acknowledgements

Authors from UNC-Chapel Hill were supported by a grant from the National Institutes of Health (Grant OT2TR002514).

## Footnotes

1 https://cloud.google.com/text-to-speech

2 This was an actual word in the text. The natural language text is discussed by non-experts, and therefore involves a lot of colloquial terms. Our language model can handle this.

